# Performance of Risk Models to Predict Mortality Risk for Patients with Heart Failure: Evaluation in an Integrated Health System

**DOI:** 10.1101/2023.06.23.23291822

**Authors:** Faraz S. Ahmad, Ted Ling Hu, Eric D. Adler, Lucia C. Petito, Ramsey M. Wehbe, Jane E. Wilcox, R. Kannan Mutharasan, Beatrice Nardone, Matevz Tadel, Barry Greenberg, Avi Yagil, Claudio Campagnari

## Abstract

**Background:** Referral of patients with heart failure (HF) who are at high mortality risk for specialist evaluation is recommended. Yet, most tools for identifying such patients are difficult to implement in electronic health record (EHR) systems.

**Objective:** To assess the performance and ease of implementation of Machine learning Assessment of RisK and EaRly mortality in Heart Failure (MARKER-HF), a machine-learning model that uses structured data that is readily available in the EHR, and compare it with two commonly-used risk scores: the Seattle Heart Failure Model (SHFM) and Meta-Analysis Global Group in Chronic (MAGGIC) Heart Failure Risk Score.

**Design:** Retrospective, cohort study

**Participants:** Data from 6,764 adults with HF were abstracted from EHRs at a large integrated health system from 1/1/10-12/31/19.

**Main Measures:** One-year survival from time of first cardiology or primary care visit was estimated using MARKER-HF, SHFM and MAGGIC. Discrimination was measured by the area under the receiver operating curve (AUC). Calibration was assessed graphically.

**Key Results:** Compared to MARKER-HF, both SHFM and MAGGIC required a considerably larger amount of data engineering and imputation to generate risk score estimates. MARKER-HF, SHFM, and MAGGIC exhibited similar discriminations with AUCs of 0.70 (0.69-0.73), 0.71 (0.69-0.72), and 0.71 (95% CI 0.70-0.73) respectively. All three scores showed good calibration across the full risk spectrum.

**Conclusions:** These findings suggest that MARKER-HF, which uses readily available clinical and lab measurements in the EHR and required less imputation and data engineering than SHFM and MAGGIC, is an easier tool to identify high-risk patients in ambulatory clinics who could benefit from referral to a HF specialist.

## Introduction

Heart failure (HF) is a heterogenous, morbid condition that affects over 6.5 million adults in the United States.^1^ Mortality risk, while variable, is considerable and approaches 50% at five years after HF diagnosis with little variation across left ventricular ejection fraction (LVEF) spectrum.^1^ This risk is not always readily apparent to the wide range of clinicians, many of whom do not have training with diagnosing and managing patients with advanced HF, who take care of these patients in various care settings. The 2022 AHA/ACC/HFSA Guideline for the Management of Heart Failure states that increased predicted one year mortality is an indicator of advanced HF and that patients with advanced HF, when consistent with a patient’s goals, is a Class I recommendation.^2^ However, referral to a HF specialist for advanced therapies evaluation is often delayed when clinicians fail to recognize the severity of a patient’s condition and to identify those at high risk of death. This may result in lost opportunities to counsel patients and their families or initiate advanced therapies evaluation for cardiac transplantation or left ventricular device implantation (LVAD), which are life-saving therapies for a subset of patients with advanced HF.^3^

The widespread adoption of electronic health records (EHRs) has created an opportunity to develop targeted health management strategies based on risk models for patients with HF.^4^ However, many HF risk models, similar to many models for other conditions, were developed relying on data from outside of health systems, such as community-based, observational cohorts or clinical trials. They frequently rely on variables that may be subjective or that are not readily available in the EHR for data analytics. In addition, many of these risk models use medication data, which is challenging to accurately extract from the EHR due to the lack of uniformity in the structure and reporting of dosing information.^5–8^

The Machine learning Assessment of RisK and EaRly mortality in Heart Failure (MARKER-HF) is an externally-validated, boosted decision tree-based machine learning model that uses eight commonly measured variables (7 laboratory measurements plus diastolic blood pressure) to estimate 1-year risk of mortality in patients with HF.^9^ MARKER-HF was developed using inpatient and outpatient EHR data from patients treated at a large academic health center. Its performance has been shown to be superior to other HF and general risk models in diverse HF populations and in HF subgroups defined by LVEF.^10^ One important strength of MARKER-HF is its use of variables that are readily available in EHR repositories. This feature makes implementation relatively straightforward in clinical settings and could facilitate execution across a broad population of patients with HF in a health system.

In this study, we compared the ease of implementation and the performance of two of the most widely used and tested HF risk models, Seattle Heart Failure Model (SHFM) and Meta-Analysis Global Group in Chronic (MAGGIC) Heart Failure Risk Score,^11–19^ with MARKER-HF in ambulatory patients with HF treated at a large, integrated health system using EHR data. We hypothesized that MARKER-HF would be easier to implement and require less data engineering than SHFM and MAGGIC while having similar overall performance for predicting 1-year risk of mortality in a larger, more representative population of patients with HF.

## Methods

The data supporting the findings of the study are available from the corresponding author upon reasonable request and a data use agreement. The Northwestern University Institutional Review Board approved this study.

### Data source and Participants

We identified a retrospective cohort of patients ages 18 to 89 years old with HF who visited outpatient primary care or cardiology at least once between 1/1/2010 to 12/31/2018 from the Northwestern Medicine Enterprise Data Warehouse (NMEDW), which houses comprehensive demographic, diagnostic, and prescription data from the 10 hospitals and over 100 sites across the integrated health system.^20^

Prevalent HF was defined by having a minimum of one inpatient or two outpatient diagnosis codes from distinct encounters for HF-based on a previously published algorithm that has been validated in the NMEDW.^21, 22^ The index visit, which was the date of prediction, was defined as either the first primary care or cardiology visit after first inpatient HF diagnosis code in the study period or the first visit of the two qualifying ambulatory visits with a HF diagnosis code. Follow-up extended through 12/31/2019.

Patients were excluded if they underwent heart transplantation or left ventricular assist implantation prior to or during the study period due to inability to accurately identify the date of surgery for the entire cohort. Patients who were not documented as deceased in the NMEDW and did not have a face-to-face encounter between one-year post-index visit through the end of the study period were excluded from the analysis. Race, ethnicity, and gender were captured as structured data in the EHR.

### Outcome

In all 3 HF risk prediction models, the outcome is death from all causes within one year of the index event (defined above). This information was captured in the NMEDW.

### HF Risk Prediction Model Inputs

Table 1 shows data availability and definitions for each variable included in the MARKER-HF, SHFM, and MAGGIC models. Additional details on the definitions and lookup period for each variable are available in Supplemental Tables S1. Many variables were available as structured data from the EHR data repository and were abstracted directly into analytic datasets; here we highlight variables identified in other manners. As previously noted, each variable incorporated into MARKER-HF is available as structured data in the EHR data repository; the following variables were used in SHFM or MAGGIC.

**Table 1:**
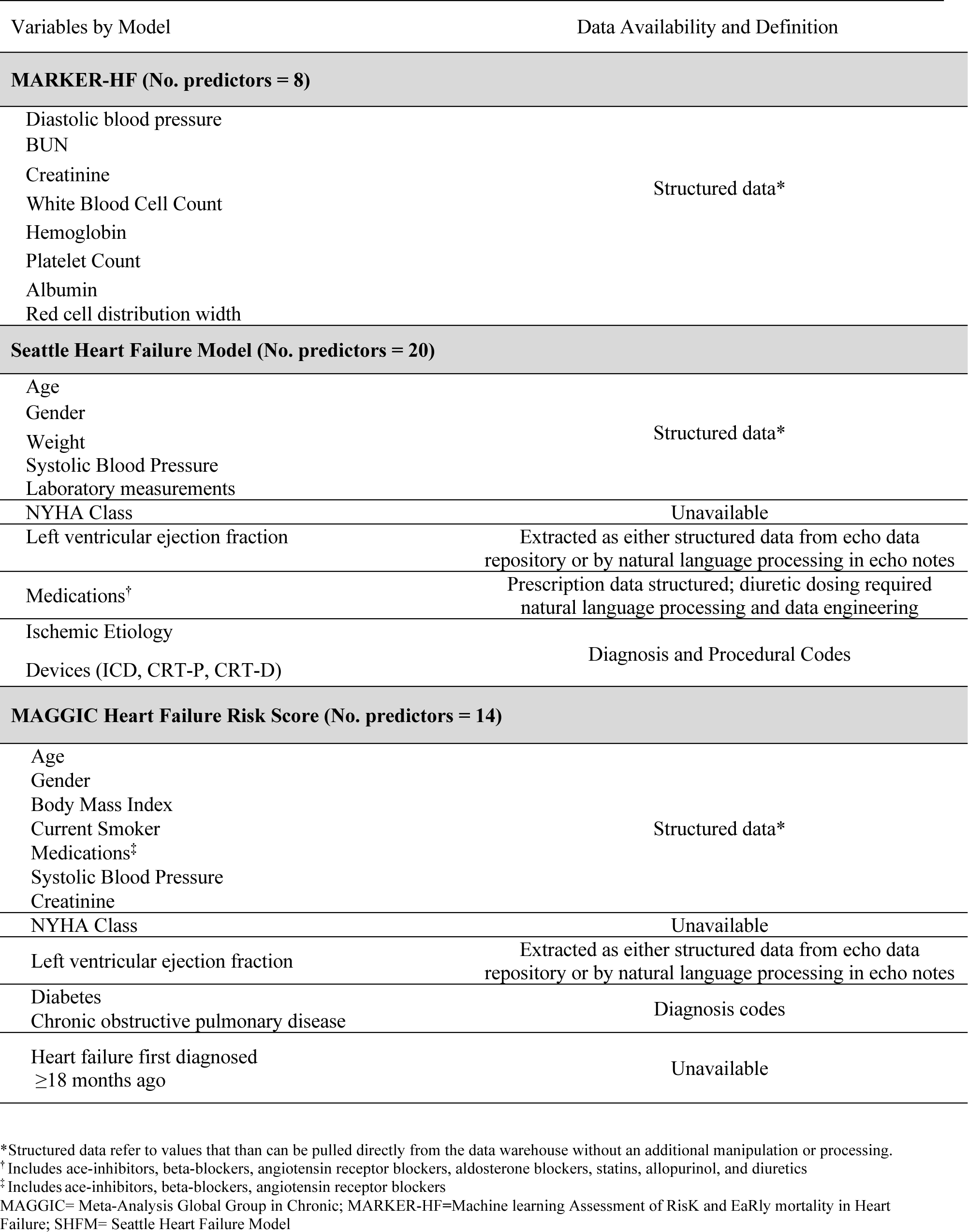
Data availability and definitions for risk predictors in MARKER-HF, Seattle Heart Failure Model, and MAGGIC Heart Failure Risk Score

We used a combination of diagnosis and procedure codes to identify patients the presence of one of three types of cardiovascular implantable electronic device (CIEDs): implantable cardioverter defibrillator, chronic resynchronization therapy pacemaker, or combined device. History of chronic obstructive pulmonary disease, diabetes, and ischemic cardiomyopathy were ascertained using diagnosis codes. Because a history of HF first diagnosed ≥18 months ago is not readily available as structured data and challenging to extract with natural language processing, all patients were considered to have a history of HF first diagnosed < 18 months. NYHA was only available in free text in a small subset of notes, so we assumed all patients had a mean value of 2.5 as previously done.^11^

### Creation of Common and Analytical Cohorts

To evaluate relative ease of implementation of MARKER-HF with SHFM and MAGGIC, we first excluded patients with insufficient follow-up time, history of heart transplant or LVAD, and EHR data quality issues. We then examined the degree of data engineering and missingness and imputation requirements in the remaining cohort (n=9,231; Figure 1).

**Figure 1:**
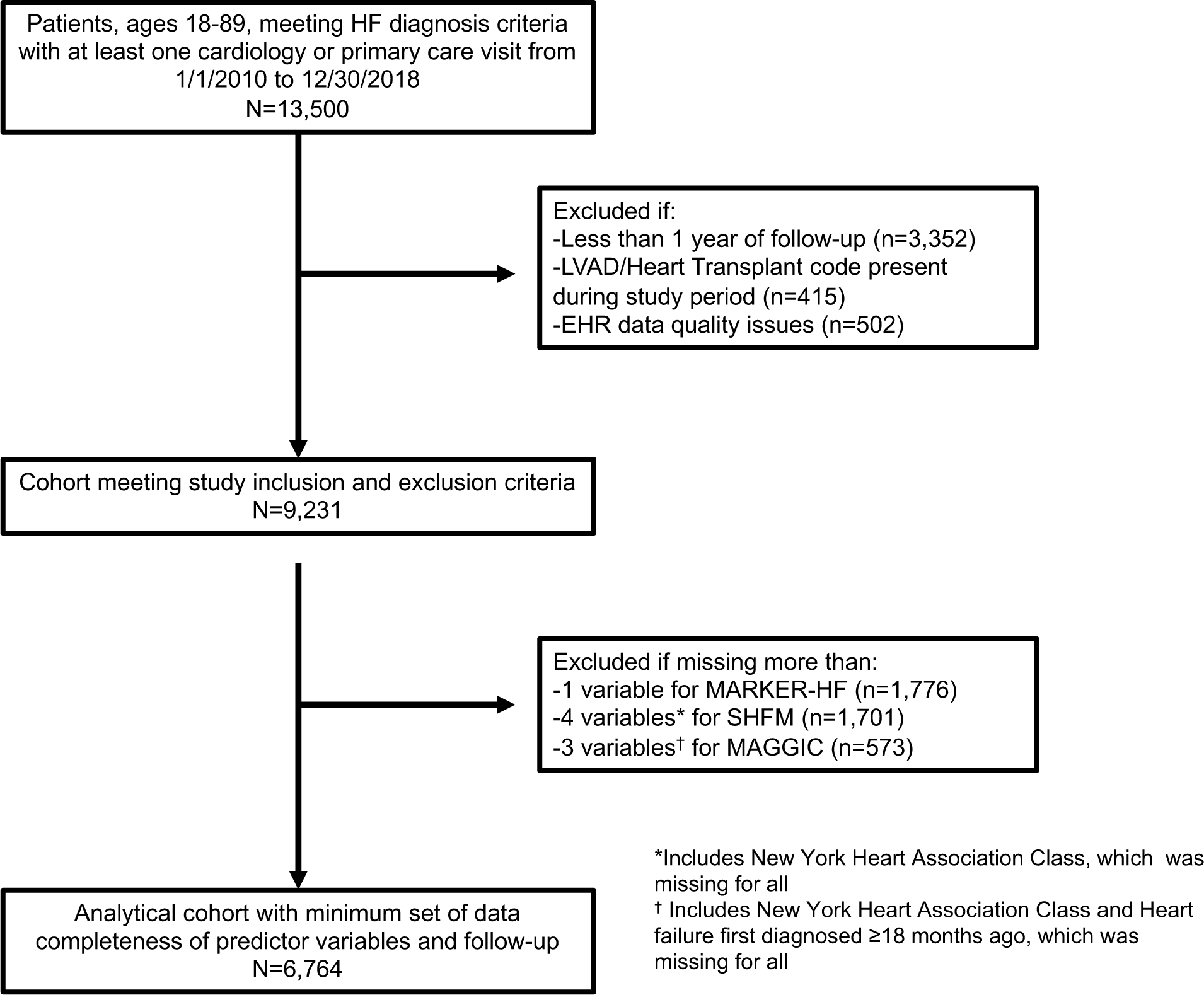
Flowchart for Cohort Identification in a Large, Regional Health System

Missing values for systolic blood pressure, weight, laboratory measurements, and LVEF, factors used in SHFM and MAGGIC, were imputed with single imputation using chained equations due to the high percentage of missing values for multiple variables. MARKER-HF score had much fewer missing values and those were imputed using mean value based on guidance from the model developers (AY and CC) and prior analysis demonstrating that the imputation of one of eight variables did not lead to substantial decrement in model performance.^9^ Timing of HF diagnosis and NYHA Class were imputed for SHFM and MAGGIC as described above.

To evaluate model performance, we then aimed to create an analytical cohort that minimized the number of imputed variables for the models while also maximizing the size of the analytic cohort (Figure 2). We therefore included patients with up to one missing variable for MARKER-HF, up to three missing for MAGGIC, and up to four missing variables for SHFM to achieve a reasonable cohort size of 6,764 patients.

**Figure 2:**
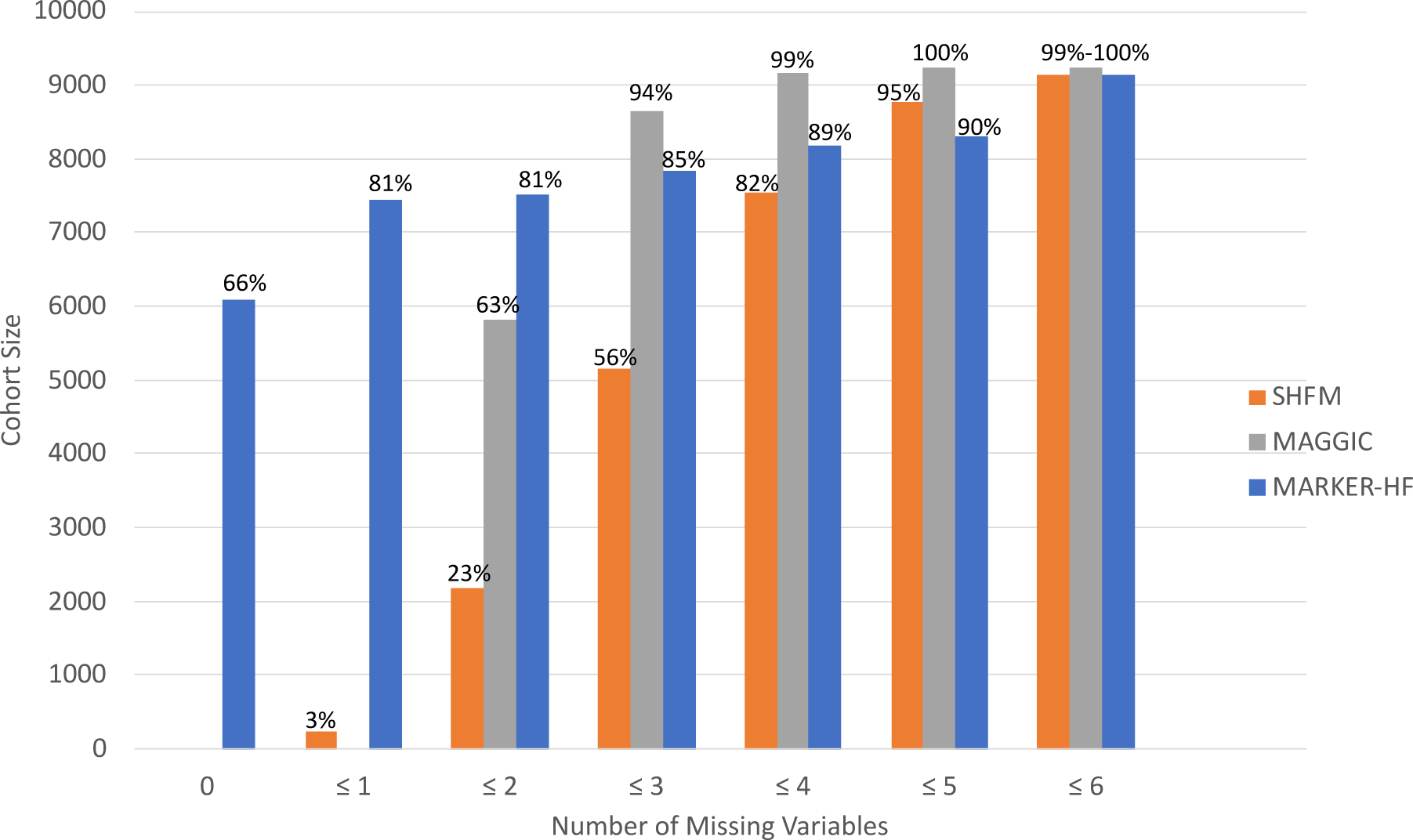
Cohort size by number of missing variables in 9,231 patients meeting study criteria. This bar chart depicts the size (percentage) of the cohort for which each score can be computed for a given number of missing variables in the cohort of 9,231 patients meeting the inclusion and exclusion criteria for the study. The percentage of top of each bar represents the number of patients in each bar divided by the cohort size (n=9,231). MARKER-HF**=**Machine learning Assessment of RisK and EaRly mortality in Heart Failure; SHFM**=** Seattle Heart Failure Model; MAGGIC= Meta-Analysis Global Group in Chronic

### Statistical Analysis: HF Risk Model Implementation and Validation

We generated MARKER-HF risk scores (range −1 to +1) and 1-year survival estimates according to the code available on GitHub at https://github.com/claudiocc1/MARKER-HF and online calculator available at https://marker-hf.ucsd.edu/. We calculated risk scores and 1-year survival estimates for the SHFM and MAGGIC using published algorithms.^13, 17^

We evaluated model discrimination, the ability of a model to correctly classify HF patients as alive or dead 1-year after the index date, using area under the receiver operating curve (AUC, or c-statistic) and compared AUCs between models with the DeLong Test.^23, 24^ We assessed model calibration, the ability of a model to closely estimate the underlying risk, by comparing observed vs predicted 1-year survival for each model.

To further evaluate model performance in subgroups, we estimated AUC in patients with each HF subtype: 1) HF with preserved EF (HFpEF) with LVEF (≥ 50%); 2) HF with mildly reduced EF (HFmrEF) with LVEF between 41% and 49%; and 3) HF with reduced EF (HFrEF) with LVEF ≤ 40%. We also estimated AUC in race and gender subgroups.

To evaluate the risk of bias from right-censoring, we compared selected baseline characteristics and risk estimates from those included in the analytical sample to those who were lost to follow-up prior to one year. In addition, we performed two sensitivity analyses. First, we evaluated the discrimination of MAGGIC in a subset of patients who almost certainly were diagnosed with HF < 18 months prior to index date: those with at least 18 months of HF diagnosis code-free data in the EHR prior to index visit and for whom time of index visit equaled time of first HF diagnosis code. Second, we evaluated the performance of SHFM and MAGGIC with the use of mean imputation instead of chained equations to mirror the imputation approach for MARKER-HF. This is likely the strategy that would be used if the models were implemented in a health system. All analyses were conducted using scikit-learn version^25^ 1.01, Lifelines^26^ version 0.26.4, and SPSS, Statistics, version 28.0.1.

## Results

### Baseline Characteristics

From the health system electronic data warehouse, we identified 13,500 patients with diagnosed HF and at least one primary and cardiology ambulatory visit between 2010 and 2018 (Figure 1). After excluding patients with insufficient follow-up time, history of heart transplant or LVAD, and EHR data quality issues, 9,231 patients were remaining in the cohort. After excluding patients with more than 3 missing variables for MAGGIC (n=573), 4 missing variables for SHFM (n=1,701), and more than 1 missing variable for MARKER-HF (n=1,776) the resulting analytical cohort had 6,764 patients remaining. In the analytical cohort, median age was 71 years (IQR 59-82), 54% were women, 90% were Non-Hispanic, and 20% were Black or African American (Table 2). This cohort had a high comorbidity burden: 40% had diabetes, 27% had chronic obstructive lung disease, and two-thirds had ischemic heart disease. 1,266 (19%) patients died within the first year of follow-up.

**Table 2:** Baseline characteristics for the analytical cohort and those who excluded due follow-up of less than one year

| **Variable** | **Analytical Cohort**<br>**(n=6,764)** | **Cohort lost to**<br>**follow-up**<br>**(n=2,295)** |
| --- | --- | --- |
| Age at time of prediction, median (IQR) | 71 (59-82) | 70 (58-80) |
| Female, n (%) | 3,627 (54) | 1,277 (56) |
| Ethnicity, n (%) |  |  |
| Hispanic | 402(6) | 168 (7) |
| Non-Hispanic | 6,068(90) | 1,888 (83) |
| Not Available | 291 (4) | 230 (10) |
| Race, n (%) |  |  |
| White | 4,601 (68) | 1,364 (60) |
| Black or African American | 1,323 (20) | 451 (20) |
| Asian | 193 (3) | 71 (3) |
| Other | 412 (6) | 185 (8) |
| Not Available | 218 (3) | 203(9) |
| Current Smoker, n (%) | 476 (7) | 176 (8) |
| Diabetes, n (%) | 2,727 (40) | 897(39) |
| Chronic Obstructive Lung Disease, n (%) | 1,815 (27) | 569(25) |
| Ischemic Heart Disease, n (%) | 4,533 (67) | 1,492 (65) |
| BMI, mean (SD) | 28 (6) | 27 (6) |
| Heart failure subtype, n (%) |  |  |
| HFrEF ($\leq 40\%$) | 1,954 (29) | 750 (33) |
| HFmrEF (41%-49%) | 529 (8) | 222 (10) |
| HFpEF ($\geq 50\%$) | 2,809 (42) | 970 (42) |
| Sodium, mean (SD), meq/L | 138 (4) | 137 (4) |
| Creatinine, mean (SD), mg/dL | 1.5 (1.4) | 1.5(1.3) |
| Albumin, mean (SD), g/dL | 3.6 (0.6) | 3.6(0.6) |
| MARKER-HF Risk Score, mean (SD) | -0.3 (0.2) | -0.2 (0.3) |
| Seattle Heart Failure Risk Score, mean (SD) | 0.97 (0.7) | 1.1 (0.7) |
| MAGGIC Heart Failure Risk Score, mean (SD) | 22.2 (6.7) | 22.2 (7.1) |

MARKER-HF=Machine learning Assessment of Risk and Early mortality in Heart Failure
MAGGIC=Meta-Analysis Global Group in Chronic
HFrEF=Heart failure with reduced ejection fraction
HFmrEF=Heart failure with mildly reduced ejection fraction
HFpEF=Heart failure with preserved ejection fraction

### Evaluation of ease of implementation and imputation requirements for each model

Of the 20 variables used to calculate the SHFM score, 9 required data engineering (i.e diuretic dosing, use of computable phenotypes, and natural language processing to extract LVEF) and 1 variable (NYHA) required 100% mean imputation since unavailable for all patients. Of the 14 variables MAGGIC uses, 5 required data engineering (i.e. use of computable phenotypes, LVEF extraction) and 2 variables (NYHA and history of HF first diagnosed ≥18 months ago) were 100% missing and required imputation. Although we employed previously published algorithms to identify medical history, computable phenotypes have varying accuracy, and ranged from using a list of codes (i.e. diabetes, chronic obstructive lung disease) to combination of codes (i.e. CIED classification). MARKER-HF did not require comparable data engineering or use of computable phenotypes. No value for MARKER-HF was 100% missing from the cohort.

Figure 2 depicts the number of missing input variables for the three scores. It includes anthropometric, diagnostic testing, and clinical (NYHA for both SHFM and MAGGIC and variables and history of HF first diagnosed ≥18 months ago for MAGGIC) for each model and the resulting cohort sizes. With imputation of one variable, MARKER-HF can be executed on 81% (7,455/9,231). For SHFM and MAGGIC, the imputation of four and three variables, respectively, were required to achieve a similar cohort size. Additional details on missingness for each variable in the analytical cohort are shown in Table S2.

### Performance of HF Risk Models

MARKER-HF and SHFM demonstrated similar model discrimination. As shown in Figure 3, the AUC for MARKER-HF (0.70; [95% CI 0.69-0.72]) was similar to SHFM (0.71; [0.69-0.73]; DeLong test P=0.64) and MAGGIC (0.71; [0.70-0.73]; DeLong test P=0.43).

**Figure 3:**
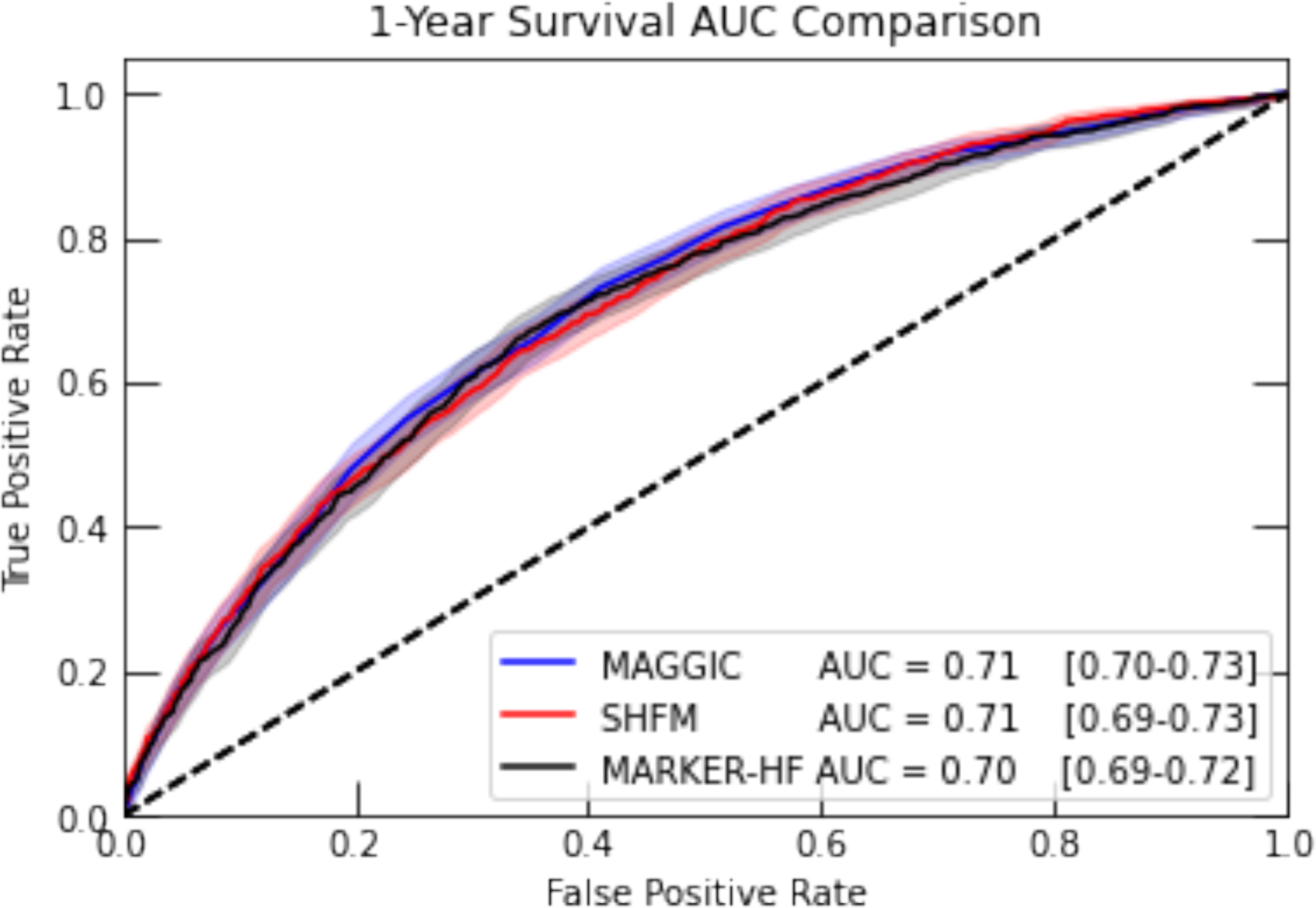
Receiver operating curve and corresponding area under the curve from 3 models (MARKER-HF, SHFM and MAGGIC) of 1-year survival in heart failure patients in a large, regional US health system. DeLong AUC test for MARKER-HF vs SHFM and MARKER-HF vs MAGGIC had p-values of 0.64 and 0.43 respectively. MARKER-HF= The Machine learning Assessment of RisK and EaRly mortality in Heart Failure; MAGGIC= Meta-Analysis Global Group in Chronic; SHFM= Seattle Heart Failure Model.

The calibration, i.e. how well predicted risks match observed risks, for MARKER-HF, SHFM, and MAGGIC was good over the full range of predicted risk (Figure 4). There was indication for over-estimation of 1-year mortality risk in the highest risk group by all three models.

**Figure 4:**
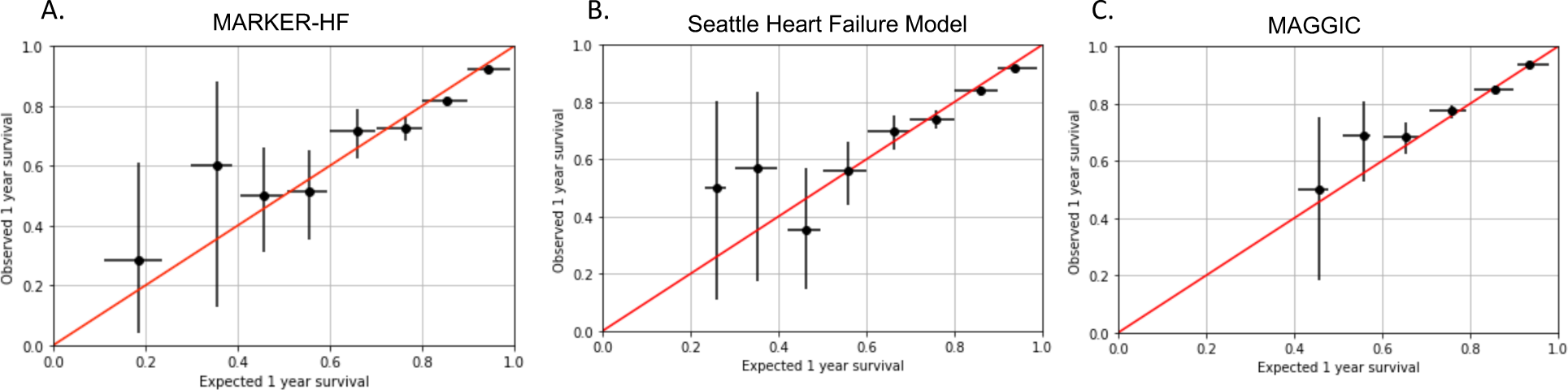
Calibration of 3 models (MARKER-HF, SHFM and MAGGIC) of 1-year survival in heart failure patients in a large, regional US health system. A) MARKER-HF; B) SHFM; and C) MAGGIC. Error bars represent the one sigma statistical uncertainty on the mean. MARKER-HF=The Machine learning Assessment of RisK and EaRly mortality in Heart Failure; MAGGIC=Meta-Analysis Global Group in Chronic (MAGGIC); SHFM=Seattle Heart Failure Model.

In subgroup analyses stratified by LVEF subtype, race (limited to patients who were Black and White due to small sample size leading to imprecise estimates in other groups), and gender, MARKER-HF, SHFM and MAGGIC all had similar performances, well within the confidence intervals for AUC from the subgroup analyses overlapping with the confidence intervals for the AUCs in the primary analysis (Supplemental Figures S1, S2, and S3). In a sensitivity analysis of MAGGIC examining a subpopulation of patients more likely HF diagnosed < 18 months at time of prediction (n=4908), the discrimination was similar (0.72; [95% CI 0.70-0.74]). In a sensitivity analysis of using mean imputation for systolic blood pressure, weight, laboratory measurements, and LVEF instead of chained equations for SHFM and MAGGIC, the change in AUC was insignificant (<5%).

## Discussion

This study used data extracted from an enterprise data warehouse to evaluate the ease of implementation and the potential value of embedding three established risk scores into the EHR and incorporating them into routine care. MARKER-HF, a model that exclusively uses information available in the EHR, required less data engineering and imputation than SHFM and MAGGIC and was easier to implement. The imputation of only 1 variable for MARKER-HF enabled the execution of the risk score on 81% of patients meeting the cohort inclusion and exclusion criteria, whereas as SHFM and MAGGIC at a minimum required imputation of 4 and 3 variables, respectively, to achieve a comparable cohort size. In the analytical cohort of 6,674 patients, MARKER-HF had similar discrimination and calibration to SHFM and MAGGIC. These findings were largely similar in subgroup analyses by HF class based on LVEF, race and gender. Results were also similar in sensitivity analyses that evaluated the potential effect of inaccurate HF diagnosis date in the EHR database on MAGGIC performance and the effect of different imputation strategies on SHFM and MAGGIC performance.

Incorporating holistic risk HF prediction tools into routine care can provide valuable insight to inform patient-clinician shared decisions as well as potential population health strategies. Yet, many systems have not adopted these tools due to workflow issues – rather, the manner in which data are stored in the EHR renders many of the variables that are required for risk-prediction tools difficult to access. For example, NYHA class, which contributes substantially to the SHFM and MAGGIC scores, must be abstracted from clinician notes, embedded in the EHR where its presence is variable. In addition to potentially leading to misclassification or inaccuracies due to imputation(s) or computable phenotype definitions, our experience in generating this comparison was that it required a considerable amount of computational effort and resources to curate and analyze the data required for calculating the SHFM and MAGGIC from the EHR.

Our analysis demonstrates that health systems do not need to undertake the additional backend work required to implement SHFM and MAGGIC; embedding MARKER-HF into the EHR could provide similarly valuable HF mortality risk information to the treating clinician or population health team. Moreover, by using readily available variables and relatively-straightforward, publicly available Python code for implementation in the production setting, MARKER-HF would likely be more straightforward to build into the EHR and maintain than more complex risk scores that require a significant amount of data engineering and imputation.

Multiple factors likely contribute to the similar model performance of MARKER-HF, compared with SHFM and MAGGIC, despite the relative simplicity of its inputs. First, MARKER-HF employs a boosted decision tree-based machine learning model, which can capture complex correlations between the inputs. Second, implementing SHFM and MAGGIC in EHR data requires imputation for outright missing information, as well as reliance on computable phenotypes based on diagnosis and procedure codes. For example, both SHFM and MAGGIC require NYHA class, which is unavailable in structured EHR data. Our imputed value of 2.5 removes the nuance introduced into the SHFM and MAGGIC scores by the extreme values. Identification of current smoking status, presence of CIED and type, etiology of HF, chronicity of HF, co-morbidities (such as diabetes and chronic obstructive pulmonary disease), and in particular medication prescription and diuretic dosing, are highly challenging to obtain reliably using EHR data.^5–8^

The concept of embedding predictive analytics in routine care to inform population health strategies and shared-decision making is central to the creation of learning health systems, which are care systems where all available data are used to enable evidence-based care equitably while also generating new evidence to inform future clinical care decisions.^27^ Using risk as part of care decision is particularly important for HF. Studies suggest that patients, particularly racial and ethnic minorities and other under-resourced populations, are often referred to a HF specialist too late.^3, 28, 29^ This delay places them at risk for worse outcomes if advanced therapies, such as cardiac transplantation or left ventricular assist device implantation, are pursued.^3^ Moreover, estimation of mortality risk may also inform discussions on therapeutics, palliative care referral, and end-of-life decision making. Although an extensive literature of risk model derivation and validation studies for HF mortality and hospitalization exist,^30–32^ MARKER-HF has the unique ability to be executed with relative simplicity and similar performance across a large, diverse HF populations in health systems due to its use of few, routinely-collected laboratory and diastolic blood pressure measurements.

This study has limitations. Due the lack of availability of several of the SHFM and MAGGIC variables as structured data in the EHR, we used imputation, natural language processing, and computable phenotypes for several variables similar to other studies that have evaluated these models using EHR-extracted, health system data. The use of these strategies, some of which were developed by our team, may have led to misclassification or biased these risk scores to the mean. However, we based our definitions, when able, on prior validation studies of SHFM and MAGGIC and on previously published electronic health data algorithms. Furthermore, only a subsample of patients had one-year follow-up, which may have biased the sample due to right censoring. However, baseline age, gender, ethnicity, race, and risk scores were similar between those with at least of year of follow-up for documented death and those without (Table 2), which suggests that our results are representative of the larger population. MARKER-HF had a lower discrimination in this study compared to the initial development and validation paper.^9^ This is may be due to a difference in prediction task; this study used 1-year mortality as the outcome whereas prior study predicted high risk (90-day mortality) vs low risk (those who did not die within 800 days) as the main outcome as well as differences in the data sources and populations. Because the guidelines for HF specifically cite elevated one-year mortality risk as a sign of advanced HF that might trigger a referral to a HF specialist and the need to standardize the model for model comparison, we used over the same time horizon of one year for all three models.^2^

The findings from this study suggest that MARKER-HF may be informative to identify patients at high risk of death, including those who may benefit from HF specialist evaluation based on expert consensus guidance to use risk as part of the referral decision making process.^3^ It also may allow the identification of low risk patients who may require less intensive resource utilization. Although some criticism has been raised at the “black box” nature of machine learning models, other experts have argued that rigorous external validation machine learning models achieves the goals of explainability.^33^ Moreover, MARKER-HF uses variables with biological relevance to and previously described association with advanced heart failure and mortality. However, future implementation studies are needed to better understand how to embed HF risk models as part of routine care to improve patient-centered outcomes and their acceptability to clinicians and patients. This includes the development of a more robust digital infrastructure and governance system to execute predictive models and evaluate model performance and its impact on clinical care longitudinally,^34–37^ and trials like The REVeAL-HF (Risk EValuation And its Impact on ClinicAL Decision Making and Outcomes in Heart Failure) trial, which tested the impact of displaying HF risk estimates to clinicians for admitted HF patients using a clinical decision support tool in a pragmatic clinical trial in a single health system.^38^ Although this trial did not show an impact on its primary endpoints, additional studies in evaluating the use of risk of death for ambulatory patients either as part clinical decision support tool or by a population health team are needed.

In summary, in this study, we found that MARKER-HF, a machine learning model that uses readily available variables, required less imputation and data engineering and had similar discrimination and calibration to SHFM and MAGGIC in a large, diverse population of patients with HF from an integrated health system. These findings indicate that MARKER-HF is a useful tool to execute in system-wide EHR data from diverse health care settings to enable patient-clinician shared decision making and population health management strategies.

## Data Availability

All data produced in the present study are available upon reasonable request to the authors and a data use agreement.

## Acknowledgements

The statements presented in this work are solely the responsibility of the author(s) and do not necessarily represent the official views of the National Institutes of Health or the American Heart Association.

