## Supplemental Tables S1-S3; Supplemental Figures S1-S3 for "Performance of Risk Models to Predict Mortality Risk for Patients with Heart Failure: Evaluation in an Integrated Health System"

**Table S1: Variable Definitions**

| Variables by Model | Definition | Time Window around index visit (prediction date) |
|---|---|---|
| **MARKER-HF** | | |
| Diastolic blood pressure<br>Blood urea nitrogen<br>Creatinine<br>White Blood Cell Count<br>Hemoglobin<br>Platelet Count<br>Albumin<br>Red cell distribution width | Extracted as structured data from the enterprise data warehouse | Closest value within 30 days of index visit. If same number of days between value before and after index date, historical value was used. |
| **Seattle Heart Failure Model** | | |
| Age | Calculated from date of birth | Age in years on date of index visit |
| Gender | Extracted as structured data from the enterprise data warehouse | n/a |
| New York Heart Association Class | Imputed mean value of 2.5 | n/a |
| Ischemic Etiology | Diagnosis and Procedural Codes based on algorithm for coronary heart disease from Roumie et al.[1] | Code present before or up to 30 days after index visit |
| Left ventricular ejection fraction | Extracted either as either structured data from echo data repository or with natural language processing from echocardiogram notes | Measurement from echocardiogram available up to 365 days before or up to 30 days after index visit. If same number of days between value before and after index date, historical value was used |
| Medications* | Active prescriptions extracted by searching for medication names for each class in medication list. Diuretic dosing required natural language processing to calculate total dose. Diuretic conversations to furosemide equivalent were: furosemide 80 mg=torsemide 40 mg=bumetamide 2 mg=metolazone 2 mg=hydrochlorothiazide 25 mg=chlorothalidone 12.5 mg=chlorothiazide 200 mg | Active outpatient prescription present within 30 days of index visit |
| Cardiovascular Implantable Electronic Device | Diagnosis and Procedural Codes and logic adapted from Hatfield et al.[2] to identify presence of device and label as ICD, CRT-D-, or CRT-P | Code present before or up to 30 days after index visit |
| Systolic Blood Pressure<br>Weight<br>Laboratory measurements | Extracted as structured data from the enterprise data warehouse | Closest value within 30 days of index visit. If same number of days between value before and after index date, historical value was used |

| **MAGGIC Heart Failure Risk Score** | | |
|---|---|---|
| Age | Calculated from date of birth | Age in years on date of index visit |
| Gender | Extracted as structured data from the enterprise data warehouse | n/a |
| New York Heart Association Class | Imputed mean value of 2.5 | n/a |
| Heart failure first diagnosed ≥18 months ago | Assigned all patients to having heart failure first diagnosed < 18 months. | n/a |
| Current Smoker | Extracted as structured data from the enterprise data warehouse | Label as "current smoker" in social history within 30 days of index visit |
| Diabetes<br>Chronic obstructive pulmonary disease | Diagnosis codes based on the Centers for Medicare and Medicaid Services Chronic Conditions Data Warehouse[3] | Code present before or up to 30 days after index visit |
| Medications[†] | Active prescriptions extracted by searching for medication names for each class in medication list | Active outpatient prescription present within 30 days of index visit |
| Left ventricular ejection fraction | Extracted either as either structured data from echo data repository or with natural language processing from echocardiogram notes | Measurement from echocardiogram available up to 365 days before or up to 30 days after index visit. If same number of days between value before and after index date, historical value was used |
| Systolic Blood Pressure<br>Body Mass Index<br>Creatinine | Extracted as structured data from the enterprise data warehouse | Closest value within 30 days of index visit. If same number of days between value before and after index date, historical value was used. |

*Includes ace-inhibitors, beta-blockers, angiotensin receptor blockers, aldosterone blockers, statins, allopurinol, and diuretics.

[†] Includes ace-inhibitors, beta-blockers, angiotensin receptor blockers

BUN = blood urea nitrogen

ICD=Implantable Cardiac Defibrillator

CRT-D=cardiac resynchronization therapy defibrillator

CRT-P=cardiac resynchronization therapy pacemaker

MAGGIC = Meta-Analysis Global Group in Chronic

MARKER-HF=Machine learning Assessment of RisK and EaRly mortality in Heart Failure

**Table S2: Number of individual anthropometric and diagnostic testing predictor variable requiring imputation in the final analytical cohort (n=6,764), ordered from largest to smallest number of imputed variables**

| | **Imputed variables, analytical cohort, N (%)** | | |
|---|---|---|---|
| **Variable*** | MARKER-HF | Seattle Heart Failure Model | MAGGIC |
| Uric acid | | 5787 (86) | |
| Total cholesterol | | 4106 (61) | |
| Lymphocytes | | 1510 (22) | |
| Left ventricular ejection fraction | | 1472 (22) | 1472 (22) |
| Albumin | 1069 (16) | | |
| Body mass index | | | 417 (6) |
| Diastolic blood pressure | 45 (0.7) | | |
| Sodium | | 71 (1) | |
| Systolic blood pressure | | 21 (0.3) | 21 (0.3) |
| Weight | | 10 (0.1) | |
| Red cell distribution width | 9 (0) | | |
| Blood urea nitrogen | 2 (0) | | |
| Creatinine | 2 (0) | | 2 (0) |
| Platelet count | 2 (0) | | |
| White blood cell count | 0 (0) | | |
| Hemoglobin | 0 (0) | 0 (0) | |

*The list of variables in table does not include imputed New York Heart Association Class for SHFM and MAGGIC, and heart failure first diagnosed ≥18 months ago needed by MAGGIC.

MARKER-HF=Machine learning Assessment of RisK and EaRly mortality in Heart Failure
MAGGIC=Meta-Analysis Global Group in Chronic
SHFM- Seattle Heart Failure Risk Model

**Figure S1: Comparison of area under the curve for predicting 1-year survival for MARKER-HF, Seattle Heart Failure Model, and MAGGIC Heart Failure Risk Score based on left ventricular ejection fraction**

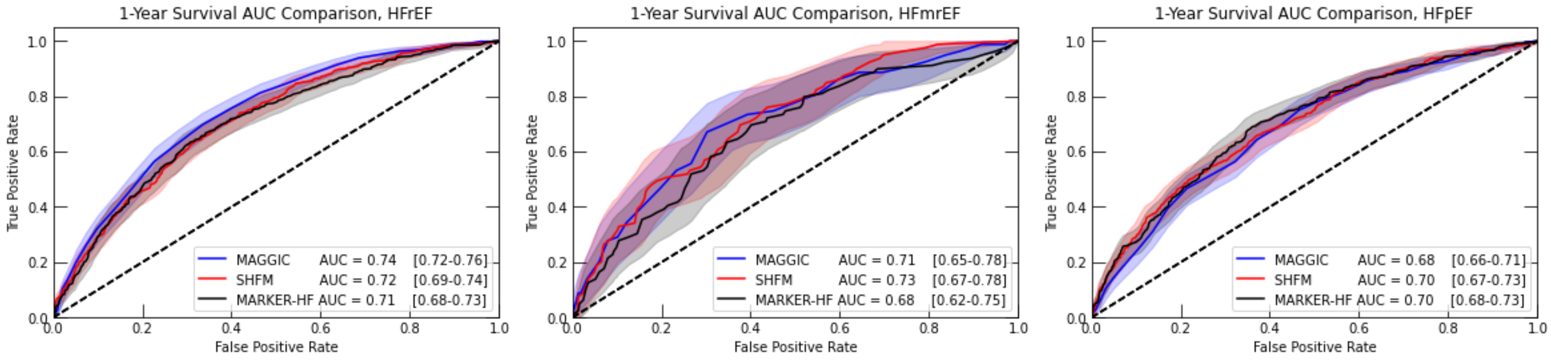


**N=3,426** **N=529** **N=2,809**

**Figure S2: Comparison of area under the curve for predicting 1-year survival for MARKER-HF, Seattle Heart Failure Model, and MAGGIC Heart Failure Risk Score for Black and White Patients with HF**

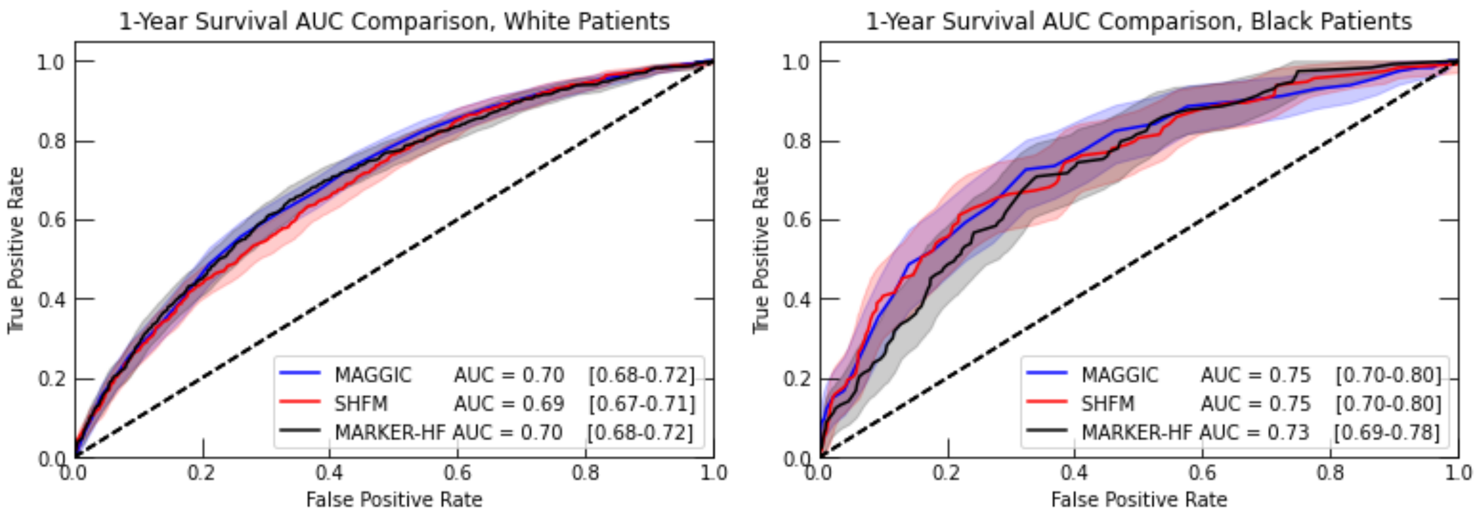


**N=4,601** **N=1,323**

**Figure S3. Comparison of area under the curve for predicting 1-year survival for MARKER-HF, Seattle Heart Failure Model, and MAGGIC Heart Failure Risk Score for Male and Female Patients with HF**

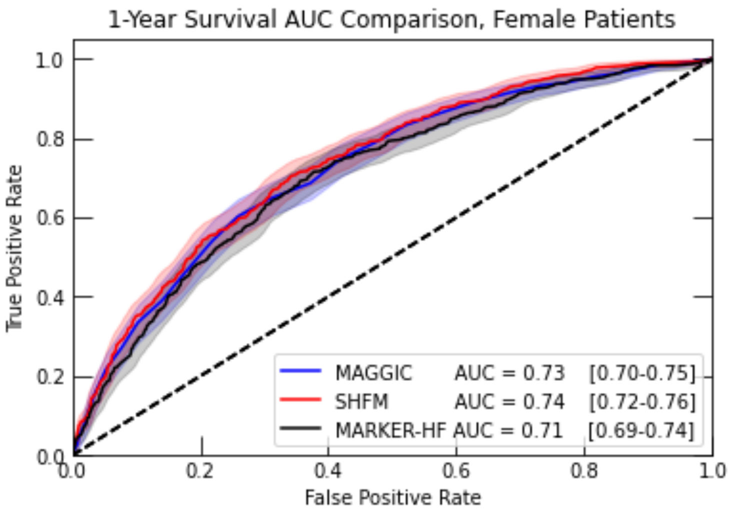


**N=3,627**

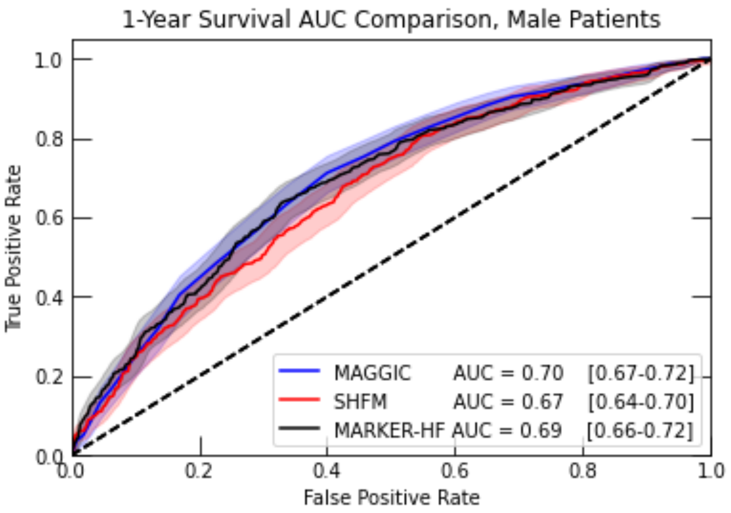


**N=3,137**
